# A Rapid and Reliable Liquid Chromatography/Mass Spectrometry Method for SARS-CoV-2 Diagnostics from Gargle Solutions and Saliva

**DOI:** 10.1101/2021.05.05.21256257

**Authors:** Marc Kipping, Dirk Tänzler, Andrea Sinz

## Abstract

We describe a rapid liquid chromatography/mass spectrometry (LC/MS) method for the direct detection and quantitation of SARS-CoV-2 nucleoprotein in gargle solutions and saliva. The method is based on a multiple-reaction monitoring (MRM) mass spectrometry approach with a total cycle time of 5 minutes per analysis and allows the detection and accurate quantitation of SARS-CoV-2 nucleoprotein as low as 500 amol/µl. We improved the sample preparation protocol of our recent piloting SARS-CoV-2 LC/MS study regarding sensitivity, reproducibility, and compatibility with a complementary reverse transcriptase quantitative polymerase chain reaction (RT-qPCR) analysis of the same sample. The aim of this work is to promote diagnostic tools that allow identifying and monitoring SARS-CoV-2 infections by LC/MS methods in a routine clinical environment.

## Introduction

The COVID-19 pandemic is one of the biggest challenges of our times. During the past 1.5 years we have seen a rapid spreading of the disease with increasing numbers of infections and fatalities. The numbers are so dynamic that it is difficult to pencil them in exact counts as are they are obsolete within hours. To date, there have been more than 140 million of SARS-CoV-2 infections and close to 3 million of fatalities worldwide,^1^ with ∼120 million of infected individuals having recovered and ∼750 million of vaccine doses administered.^1^ An infection with SARS-CoV-2 does not necessarily lead to serious health problems; often, the patients do not experience any symptoms. This is one of the greatest challenges when dealing with SARS-CoV-2, making it one of the most contagious and successfully spreading viruses. Therefore, reliable diagnostic tools allowing a rapid detection of SARS-CoV-2 are of enormous importance. Currently, the gold standard for SARS-CoV-2 diagnostics is reverse transcription quantitative polymerase chain reaction (RT-qPCR). Despite its strengths, RT-qPCR also suffers a number of drawbacks. ^2^ At the beginning of the COVID-19 pandemic in early 2020, the biggest challenge was the availability of high-throughput tools to diagnose a SARS-CoV-2 infection. This has been successfully addressed by the development of cost-efficient, ready-to-use antigen detection tests, which have become broadly available.^3^ Another important issue that needs to be dealt with is the occurrence of SARS-CoV-2 mutations that have to be detected in routine diagnostics.^4^

Mass spectrometry (MS) has already been spotted in April 2020 as a valuable complementary method for SARS-CoV-2 diagnostics from gargle solutions of COVID-19 patients.^5^ In the meantime, the potential of MS has become evident ^6^ and numerous reports on SARS-CoV-2 identification by MS methods have been published since the beginning of the pandemic.^7-11^ In the present work, we describe our latest efforts in developing a rapid liquid chromatography (LC)/MS method for SARS-CoV-2 diagnostics from gargle solutions and saliva. We report an MS-compatible sample preparation strategy and a quantification protocol that is based on selected peptides from SARS-CoV-2 nucleoprotein by an LC/multiple-reaction monitoring (MRM)-MS approach.

### Experimental Section

#### Sample collection and preparation

For LC/MS method development, two sample types - gargle solutions and saliva - were employed. Gargle solutions were collected from six healthy individuals by gargling with 20 ml of isotonic (0.9 %) NaCl solution for 30 seconds. Saliva samples were directly collected from three healthy individuals by spitting ∼1 ml of saliva into a 50-ml tube. In initial experiments, our original sample preparation procedure ^5^ was employed, adding 1 ml of acetone (−20°C) to 750 µl of gargle solution. After storing the samples at -20°C overnight, samples were centrifuged (14,000*g*, 10 min), and the protein pellets were digested using the SMART Digest (Thermo Fisher Scientific) protocol with immobilized trypsin beads. In further experiments, we optimized sample preparation in respect to shorter preparation times, higher sensitivity, and improved compatibility with RT-qPCR by using TRIZOL reagent (100 ml of solution contain 9.5 g guanidinium thiocyanate, 3.1 g ammonium thiocyanate, 3.5 ml 3M sodium acetate, 5g glycerol, 48 ml Roti Aqua-Phenol (Roth)) for protein denaturation. 500 µl of TRIZOL reagent was added to 100 µl of saliva and the solutions were loaded on centrifugation filter units (Amicon, 30 kDa molecular weight cut-off, Millipore). Afterwards, the filter units were treated according to the following filter-aided sample preparation (FASP) protocol: Filter units were washed twice with 500 µl of 50 mM ammonium bicarbonate, followed by centrifugation (14,000*g*, 10 min) and incubated at 55°C for 2 hours with trypsin (Promega) (1µg in 50 µl of 50 mM ammonium bicarbonate). Tryptic peptides were collected by adding 25 µl of 0.5M NaCl, followed by a centrifugation step (14,000*g*, 10 min). After adding 2.5 µl of 10% (v/v) trifluoroacetic acid (TFA), sample volumes were adjusted to 70 µl, before the samples were subjected to LC/MS analysis.

#### Liquid chromatography/mass spectrometry (LC/MS)

Four selected synthetic peptides derived from SARS-CoV-2 nucleoprotein (Arg41-Lsy61, Arg41-Lys65, Met210-Arg226 and Met210-Lys233) and their heavy isotope (^13^C_6_ and ^15^N_4_)-labeled variants (Arg41-Lsy*61, Arg41-Lys*65, Met210-Arg*226 and Met210-Lys*233, SpikeTide-TQL peptides, where * denotes [^13^C ^15^N ]-Arg or [^13^C_6_ ^15^N_2_ ]-Lys) were purchased from JPT Peptide Technologies and used for method development and as quantitation standards. LC separation of peptides was performed on a UPLC I-Class FTN system (Waters) equipped with a BEH C18 column (2.1 mm x 50 mm, 1.7µm, Waters). The UPLC system was directly coupled to a Xevo TQ-XS mass spectrometer (Waters) equipped with electrospray ionization (ESI) source. MS acquisition was performed using an MRM method of two selected transitions per peptide with optimized collision energies.

#### Experimental setup to estimate LOD/LOQ

Isotope-labeled peptides (see above) were diluted in 50 mM ammonium bicarbonate to concentrations between 5 amol/µl and 500 fmol/µl and digested with trypsin. Isotope-labeled peptides were spiked at different concentrations (0.5 to 50 fmol/µl) to gargle solution samples after tryptic digestion (SMART digestion protocol). Alternatively, isotope-labeled peptides were spiked at the same concentrations (0.5 to 50 fmol/µl) to saliva samples after tryptic digestion (TRIZOL/FASP protocol). The obtained results served to determine the limit of detection (LOD) and the limit of quantitation (LOQ) of our LC/MS method.

## Results and Discussion

### Optimization of LC/MS method

The aim of our LC/MS method development was the detection of four selected peptides from SARS-CoV-2 nucleoprotein that had been identified in gargle solutions of COVID-19 patients in our previous study. ^5^ We sought to achieve the highest sensitivity possible, together with short run times in a highly robust setup that might be used in a clinical routine environment. First, we had to switch from our initial nano-HPLC setup (300 nl/min) with an LC time of more than three hours ^5^ to a normal-flow UPLC setup. The most important constraint was to achieve a baseline LC separation of the four peptides from SARS-CoV-2 nucleoprotein that had been identified previously in gargle samples.^5^ For this, we applied the LC gradient as displayed in Figure 1 with a total cycle time of 5 min. It should be noted that short LC/MS cycle times are crucial for conducting medium-to high-throughput analyses in MS-based SARS-CoV-2 routine diagnostics. While sample preparation can be parallelized enabling hundreds of samples to be processed at the same time, sample throughput by LC/MS relies on short cycle times and the number of available mass spectrometers. Consequently, LC/MS cycle times of 5 minutes allow 12 samples per hour (288 samples per day) to be analyzed on each instrument. Another important aspect to be considered for LC/MS analysis in a clinical setting is a low sample carry-over from injection to injection. Carry-over between samples was reduced in our setup to non-detectable amounts by using 0.2% TFA as purge solution and 0.2% formic acid plus 0.2% TFA in water/acetonitrile (50:50 (v/v)) as washing solution of the UPLC system (data not shown).

**Figure 1:**
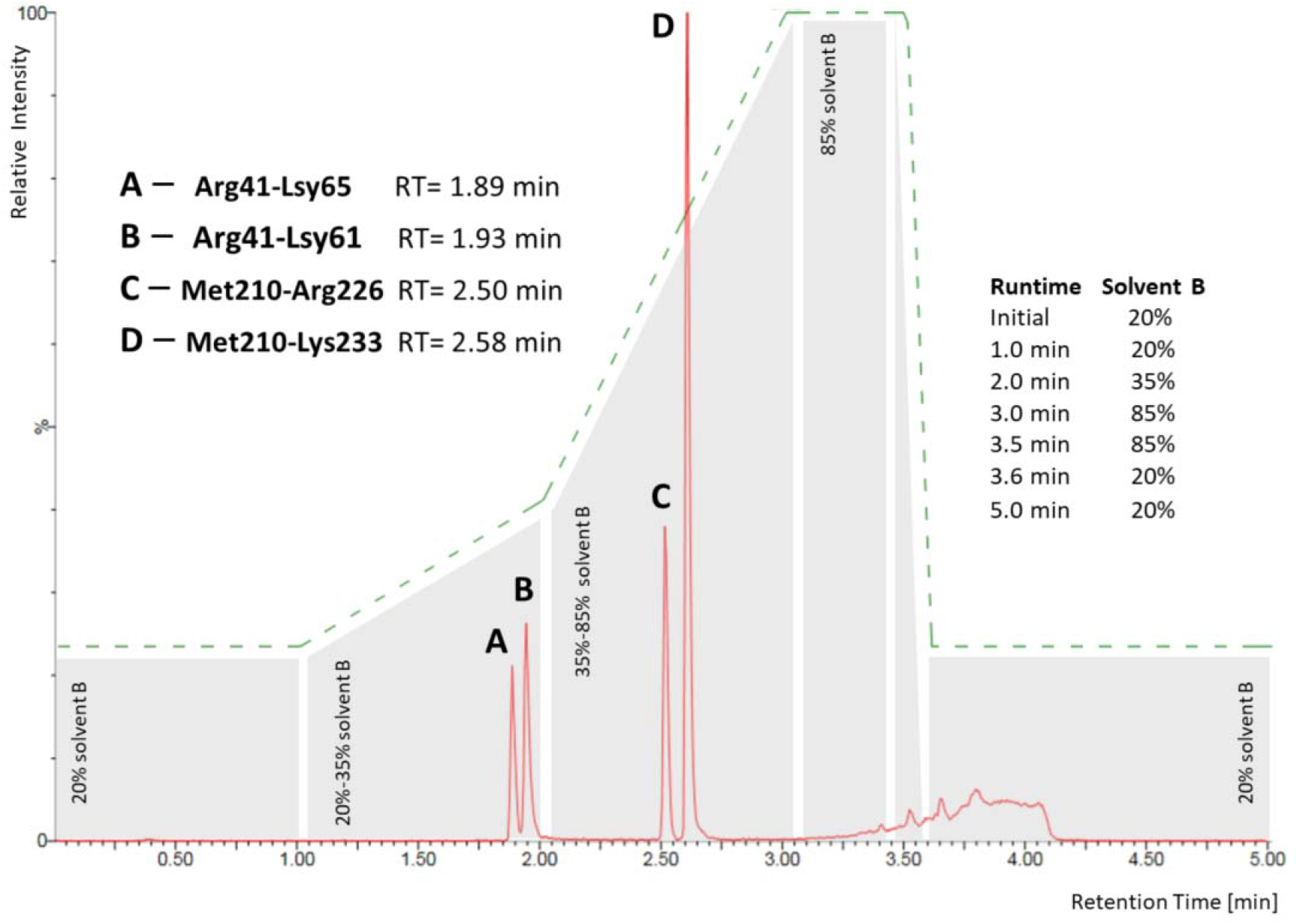
LC/MRM chromatogram of the four peptides (A) Arg41-Lsy61, (B) Arg41-Lys65, (C) Met210-Arg226, and (D) Met210-Lys233 derived from SARS-CoV-2 nucleoprotein. Peptides A-D were separated at a flow rate of 400 µl/min with the LC gradient as indicated; solvent A: 0.2% formic acid in water, solvent B: 0.2% formic acid in acetonitrile. RT: retention time.

Next, we had to move from the high-resolution mass spectrometer (Orbitrap Fusion Tribrid) that had been employed in our previous study ^5^ to a quadrupole instrument of highest sensitivity. Sensitivity is one of the key points for MS-based, reliable SARS-CoV-2 diagnostic methods as viral proteins are directly detected. This stands in contrast to RT-qPCR that relies on the amplification of the genetic material of the virus. Therefore, the detection limits for SARS-CoV-2 are directly related to the sensitivity of the LC/MS method. The most sensitive method using triple-quadrupole mass spectrometers is MRM scanning. MRM is a targeted method that has to be specifically optimized for each analyte by evaluating the optimum conditions for collisional activation and thereby identifying the most abundant and characteristic fragment ions.

As outlined above, optimizing our LC/MS method was based on synthetic variants of four peptides derived from SARS-CoV-2 nucleoprotein identified in our previous study ^5^. For these four peptides, we identified the most suitable MRM transitions and collision energies with two transitions per peptide (Table 1). We also added MRM transitions of selected tryptic peptides from other abundant proteins present in our samples to monitor them during sample preparation optimization (see below).

**Table 1:**
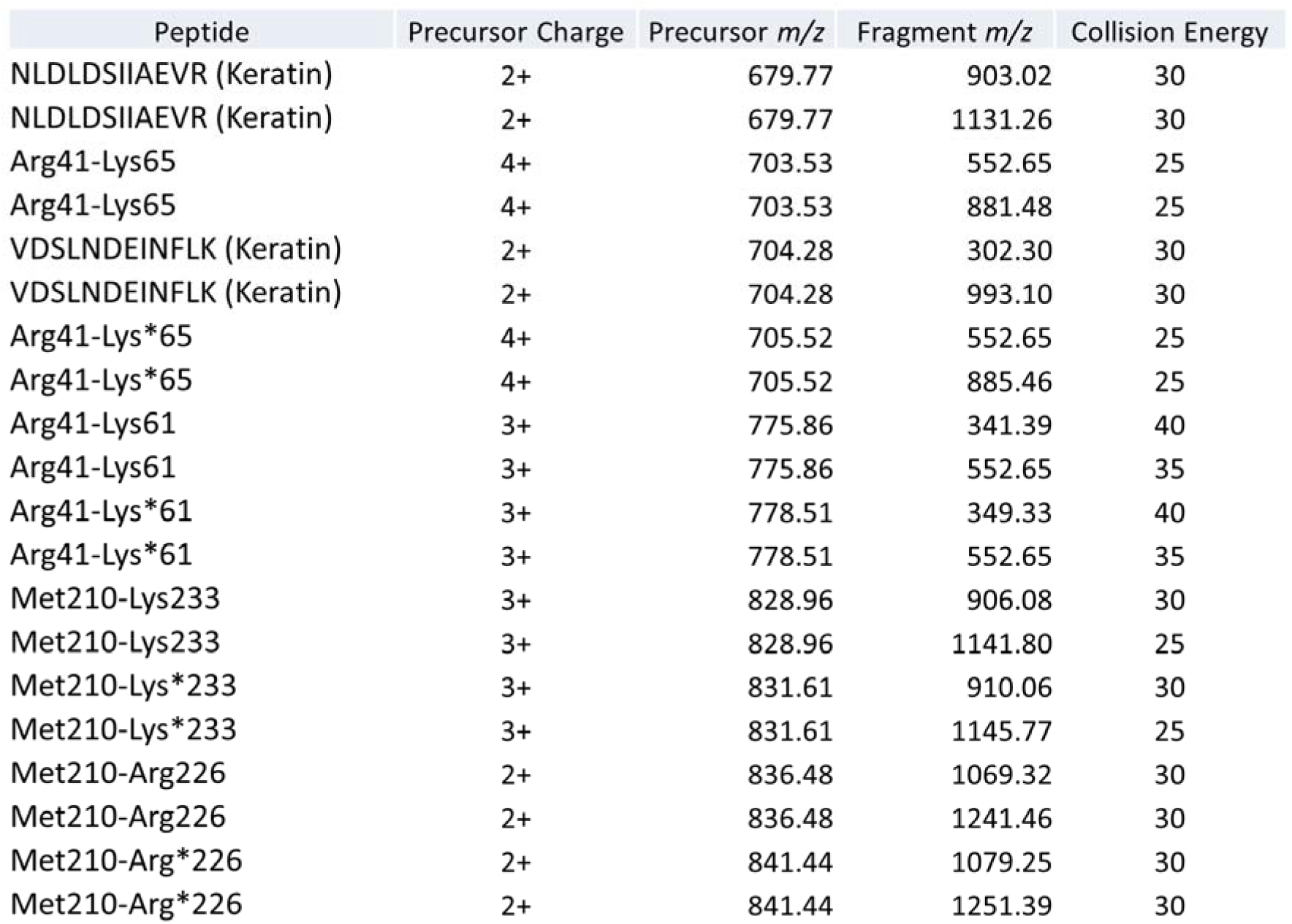
MRM transitions and collision energies of SARS-CoV-2 peptides, isotope-labeled peptides (*), and selected keratin peptides. Two transitions were selected per peptide.

All four synthetic SARS-CoV-2 peptides were employed as heavy isotope (^13^C and ^15^N)-labeled versions (SpikeTide-TQL peptides) that were added to the sample solutions for an accurate detection and quantification of SARS-CoV-2 peptides. Heavy isotope-labeled peptides only differ in the masses of precursor and fragment ions, but yield identical fragmentation patterns as the non-labeled SARS-CoV-2 peptides upon collisional activation in the MRM approach. Also, the chromatographic behavior of non-labeled and labeled peptides is identical. Therefore, MRM chromatograms of the synthetic heavy isotope-labeled peptides cannot only be used for the absolute quantification of SARS-CoV-2 nucleoprotein in patient samples, but will also exclude the assignment of false positives in highly complex matrices.

#### Sensitivity of LC/MS method and estimation of LOD/LOQ

In further experiments we then used our established LC/MS method for optimizing sample preparation. The peptides from SARS-CoV-2 nucleoprotein that had been identified in our previous study ^5^ comprise amino acid sequences Arg41-Lys65 and Met210-Lys233. These peptides contain one missed cleavage site each as our initial digestion procedure was apparently incomplete. The respective SARS-CoV-2 nucleoprotein peptides without missed cleavage sites are composed of amino acid sequences Arg41-Lsy61 and Met210-Arg226. The optimized sample preparation (see below) now allows a complete enzymatic digestion, yielding peptides Arg41-Lsy61 and Met210-Arg226 from SARS-CoV-2 nucleoprotein.

For that reason, we only employed isotope-labeled SpikeTide-TQL peptides Arg41-Lsy*61 and Met210-Arg*226 in all following experiments. SpikeTide-TQL peptides are extended by a *C*-terminal tag for enhanced solubility and photometric quantitation that is removed during tryptic digestion. It is not possible to directly add these peptides to gargle solutions or saliva as the peptides do not precipitate and will not be retained using centrifugation filter devices with a molecular weight cut-off value of 30 kDa. Therefore, isotope-labeled peptides were added after tryptic digestion in all experiments.

Figure 2A displays the LOD and LOQ of Arg41-Lsy*61 and Met210-Arg*226 isotope-labeled peptides (see Experimental Section) that had been digested with trypsin in 50 mM ammonium bicarbonate without adding protein matrix. Both LOD and LOQ were determined to be lower than 500 amol/µl. In further experiments, isotope-labeled peptides were spiked to protein pellets of gargle solutions before the SMART digestion protocol ^5^ was applied. Figure 2B displays a dramatic decrease in sensitivity and reproducibility for the SARS-CoV-2 peptides upon adding a protein matrix, underlining the need to optimize sample preparation.

**Figure 2:**
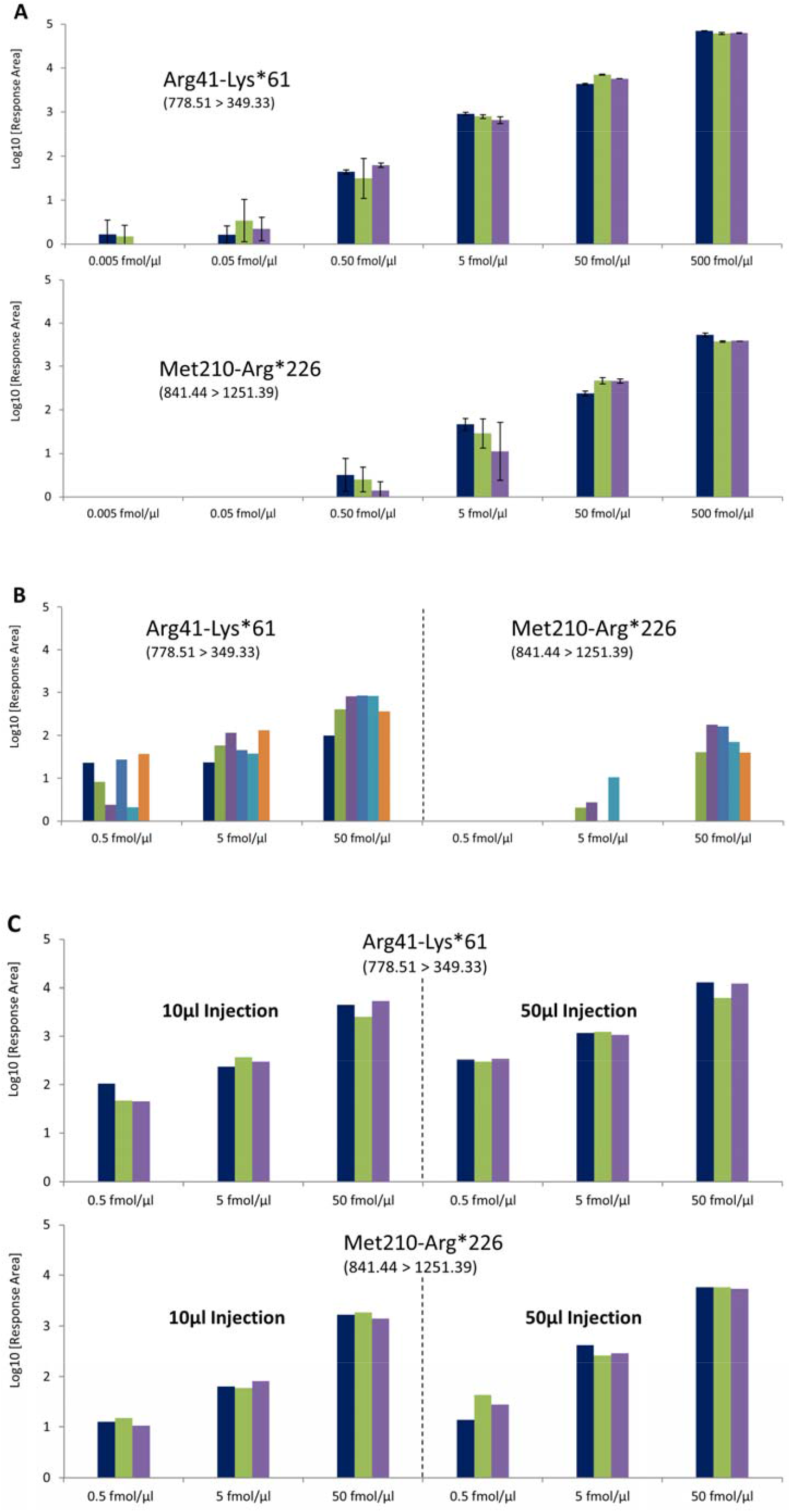
MRM detection of isotope-labeled SARS-CoV-2 peptides Arg41-Lys*61 and Met210-Arg*226 under different conditions. The observed transitions are indicated. **(A)** Dilution series of peptides in ammonium bicarbonate buffer without adding protein matrix. All experiments were performed in three replicates each (sample preparation and LC/MS measurements). **(B)** Peptides were added at three concentrations to gargle solutions, SMART digestion protocol was applied. **(C)** Peptides were added at three concentrations to saliva samples, TRIZOL/FASP protocol was applied.

#### Optimization of sample preparation

Improving our sample preparation protocol included sample collection, sample homogeneity, sample concentration, and sample preparation times. In principle, nasopharyngeal swabs, gargle solutions, and saliva are suited to detect SARS-CoV-2.^5,11-13^ A number of challenges regarding non-MS-compatible contaminations have been reported for swab samples ^11^ where the sample volume is usually less than 10 µl. Additionally, the quantitative reproducibility of swab sample collection is a matter of debate ^13^, making gargle solutions a true alternative for sample collection instead of the commonly used nasopharyngeal swab samples. Saliva of COVID-19 patients should contain even higher concentrations of SARS-CoV-2 particles than gargle solution and as saliva is easy to collect, sample quality should be highly reproducible.

Conclusively, we considered saliva to be the optimum sample for a reproducible and quantitative analysis of SARS-CoV-2 and focused our sample preparation optimization on saliva samples. First, 100 µl of saliva was treated with TRIZOL reagent for a complete dissolution of all sample components. The TRIZOL reagent is typically used for RNA preparation ^14,15^ and presents one of the most potent protein denaturants. TRIZOL reagent allows a rapid and complete inactivation of RNases, therefore enabling the isolation of intact RNA from cell lysates. Treating our samples with TRIZOL reagent leads to homogenous solutions and is therefore compatible with our FASP protocol for efficient washing and tryptic digestion of proteins. The pellet originating from acetone precipitation, as used in our previous study,^5^ was sometimes found to be incompletely dissolved in SMART digest buffer. Further advantages of the TRIZOL treatment are the direct availability of the sample as no overnight protein precipitation step is needed, and the possibility to split samples for parallel analyses by LC/MS and RT-qPCR. The TRIZOL/FASP protocol is compatible with adding the isotope-labeled peptides after tryptic digestion has been performed. Strikingly, the observed LOD/LOQ values (Figure 2C) were comparable to values obtained without protein matrix (Figure 2A).

One initial finding when optimizing the TRIZOL/FASP protocol was the detection of high amounts of cytoskeletal keratins in a number of saliva samples. Therefore, we added four specific MRM transitions for high-abundant keratin peptides (Table 1) and monitored their detection. Apparently, keratins contained in the saliva samples were efficiently removed by a simple centrifugation step before applying the FASP protocol and tryptic digestion. We speculate that without centrifugation, the TRIZOL reagent completely dissolves the keratin-containing cells in the saliva making them accessible for a subsequent tryptic digestion. This additional simple centrifugation step of saliva samples will not interfere with the overall sample preparation procedure. Nevertheless, there is a slight chance that virus particles might be attached to cells contained in saliva, which will then be removed by centrifugation. For that reason, we tested the influence of keratins on the detection of SARS-CoV-2 nucleoprotein peptides. Interestingly, neither LOD nor LOQ values were negatively affected by the presence of high amounts of keratin peptides in the samples (Figure 3). On the other hand, loading very low protein amounts (below 1µg) on the centrifugation filter devices resulted in a loss of spiked SARS-CoV-2 peptides, which is probably caused by protein and peptide binding to the membrane of the filter units. A minimum protein load of 1 µg per filter unit was obtained from treating 100 µl of saliva with TRIZOL. Additionally, sample loading might be increased to further improve sensitivity, albeit at the cost of longer sample preparation times. It is important to note that neither sample reduction, nor alkylation steps were performed in our digestion protocol as the respective SARS-CoV-2 peptides do not contain cysteine residues and proteolytic accessibility of these peptides is not restricted by disulfide bonds. In conclusion, the overall time for sample preparation can be reduced to less than three hours and can be performed in a highly parallelized fashion. Therefore, the direct use of saliva in combination with our optimized TRIZOL/FASP protocol should improve both sample collection as well as sensitivity of SARS-CoV-2 detection.

**Figure 3:**
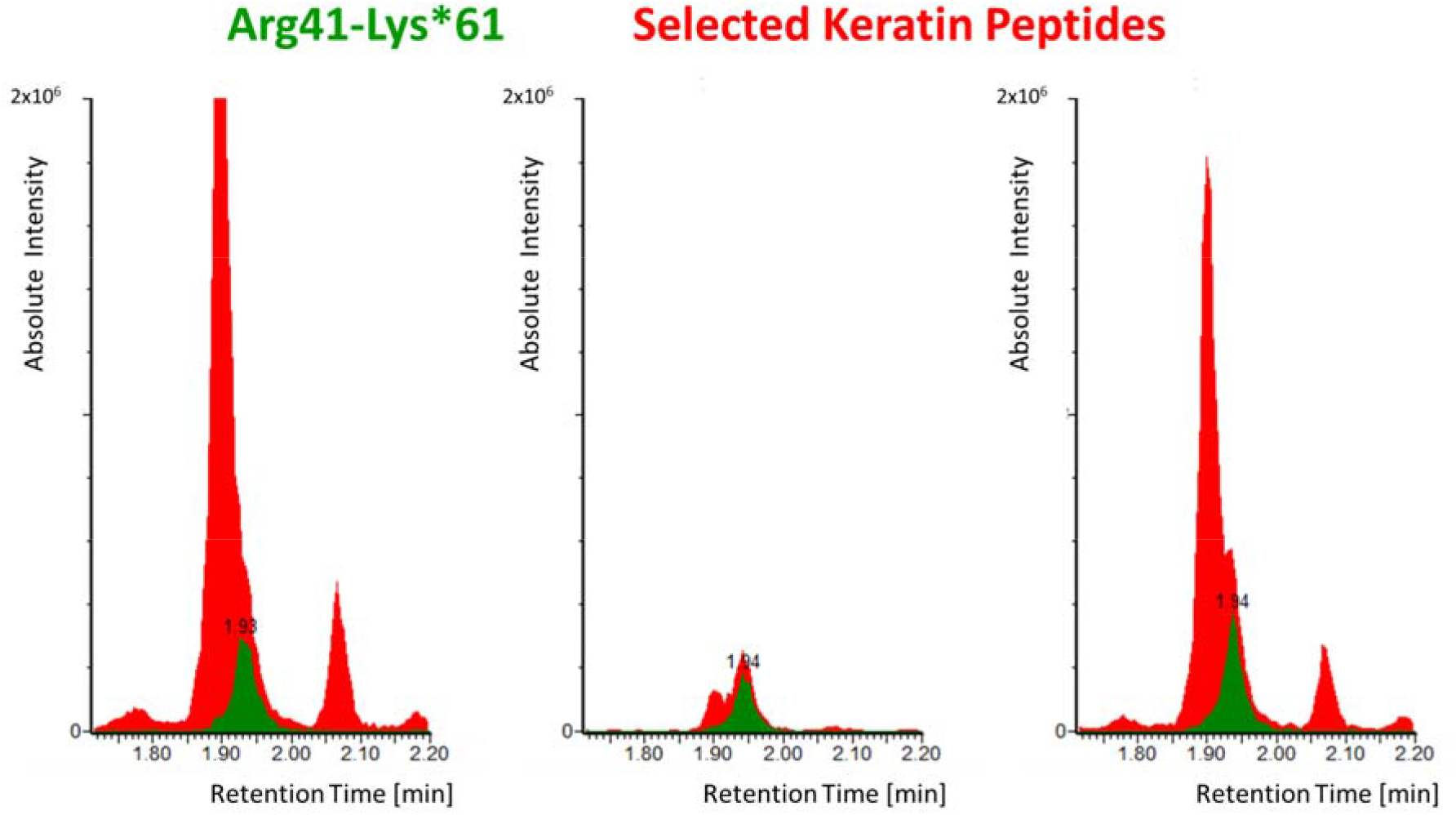
LC/MRM-MS detection of isotope-labeled SARS-CoV-2 peptide Arg41-Lys*61 at 50 fmol/µl (green) in the presence of keratin peptides (red). Selected transitions are presented: 778.51 → 349.33 (Arg41-Lys*61), 704.28 → 302.30 (keratin peptide VDSLNDEINFLK), and 679.77 → 902.02 (keratin peptide NLDLDSIIAEVR). The signal intensity of the SARS-CoV-2 peptide is not suppressed by the keratin background.

#### Comparison with RT-qPCR

It has been demonstrated that 10 genomes, i.e. DNA plasmids, ^11^ can be detected in a volume of 10 µl with cycle times (ct) of 38 in RT-qPCR experiments. A RT-qPCR detection with a ct value of 18 would require the presence of 10,485,760 genome equivalents in 10 µl. Calculations with 300 molecules of nucleoprotein per SARS-CoV-2 virus particle ^11^ result in a nucleoprotein concentration of ∼ 520 amol/µl, corresponding to a ct value of 18. ^11^ In our experiments, we used 100 µl of saliva and generated 70 µl of sample solution for LC/MS analysis. Under these conditions, the concentration of SARS-CoV-2 nucleoprotein peptides would be 743 amol/µl, which is above our estimated LOD/LOQ of ∼ 500 amol/µl (see above).

It is however very likely that a “real-world” RT-qPCR experiment is less sensitive than the one performed under the ideal conditions described in ^11^ as RNA recovery is rarely complete. Therefore, RT-qPCR experiments with specific ct values would be based on higher virus numbers, resulting in *de facto* higher nucleoprotein concentrations. The sensitivity of our LC/MRM-MS approach is likely to be further improved by using higher sample amounts, i.e. 200 or 400 µl of saliva, and by enriching the tryptic SARS-CoV-2 peptides during sample preparation.

## Conclusions

We successfully developed an LC/MS method based on an MRM approach that can be used for the direct detection of SARS-CoV-2 from saliva samples. Our approach is complementary to RT-qPCR methods and existing antigen tests, and can be used in a routine clinical environment where quadrupole mass spectrometers are available. The total analysis time of our method is five minutes, allowing SARS-CoV-2 peptides to be detected and quantified at concentrations as low as 500 amol/µl. In general, LC/MS methods exhibit improved quantitation accuracy than RT-qPCR as no amplification (10^6^-to 10^9^-times) of the analytes is involved. It is still a matter of debate how different sample collection protocols influence SARS-CoV-2 detection. MRM-MS-based methods could fill this gap and allow delivering more precise and accurate quantitative data for a direct comparison between different methods.

The outstanding advantage of LC/MRM-MS is its potential to detect SARS-CoV2 mutations. The vast majority of amino acid exchanges will result in mass shifts that are easily detected by MS, but there will probably be no drastic change in physicochemical properties, like ionization efficiency and fragmentation patterns, of the respective SARS-CoV-2 peptides. In cases virus mutations will be detected during MS analyses, novel SARS-CoV-2 standard peptides can readily be synthesized.

Conclusively, we consider the development of novel LC/MS methods to be of outstanding importance to tackle some of the most urgent issues of the COVID-19 pandemic. It seems out of question that fighting this pandemic urgently requires interdisciplinary collaborations between virologists, clinicians, and analytical chemists.

## Data Availability

All data referred to in the manuscript are in the Supplementary Informaion.

## Abbreviations

ACN: Acetonitrile
ct: Cycle time
ESI: Electrospray ionization
FA: Formic acid
FASP: Filter-aided sample preparation
HPLC: High-performance liquid chromatography
LC: Liquid chromatography
LOD: Limit of detection
LOQ: Limit of quantitation
MRM: Multiple-reaction monitoring
MS: Mass spectrometry
MS/MS: Tandem mass spectrometry
RT-qPCR: Reverse transcription quantitative polymerase chain reaction
TFA: Trifluoroacetic acid
UPLC: Ultra high-performance liquid chromatography

## >ASSOCIATED CONTENT

### Supporting Information

The Supporting Information (SARS_CoV_2_LC_MRM.xlsx) contains response areas and peak heights extracted from LC/MRM raw files (TargetLynx, Waters) as well as calculations and a complete list of MRM transitions.

## ACKNOWLEDGEMENTS

A.S. acknowledges financial support by the DFG (RTG 2467, project number 391498659 “Intrinsically Disordered Proteins – Molecular Principles, Cellular Functions, and Diseases”), the region of Saxony-Anhalt, and the Martin Luther University Halle-Wittenberg (Center for Structural Mass Spectrometry). The Waters Corporation is gratefully acknowledged for providing the Xevo TQ-XS instrument and Dr. Mike van Oosterhout, Dr. Hans Vissers, and Gunnar Weibchen for continuous support. The authors would like to thank Xiaohan Wang for excellent technical support and Dr. Christian Ihling for fruitful discussions and additional LC/MS measurements. Prof. Dr. Mike Schutkowski is acknowledged for giving access to the I-Class UPLC system to conduct initial LC/MS experiments.

## Author Contributions

All authors have given approval to the final version of the manuscript.

